# Emerging and Continuing Trends in Opioid Overdose Decedent Characteristics during COVID-19

**DOI:** 10.1101/2021.07.28.21261276

**Authors:** Gian-Gabriel P. Garcia, Erin J. Stringfellow, Catherine DiGennaro, Nicole Poellinger, Jaden Wood, Sarah Wakeman, Mohammad S. Jalali

## Abstract

**Background:** Since COVID-19 erupted in the United States, little is known about how state-level opioid overdose trends and decedent characteristics have varied throughout the country.

**Objective:** Investigate changes in annual overdose death rates, substances involved, and decedent demographics in opioid overdose deaths across nine states; assess whether 2019-2020 trends were emerging (i.e., change from 2019-2020 was non-existent from 2018-2019) or continuing (i.e., change from 2019-2020 existed from 2018-2019).

**Design:** Cross-sectional study using vital statistics data to conduct a retrospective analysis comparing 2020 to 2019 and 2019 to 2018 across nine states.

**Setting:** Alaska, Colorado, Connecticut, Indiana, Massachusetts, North Carolina, Rhode Island, Utah, and Wyoming.

**Participants:** Opioid-related overdose deaths in 2018, 2019, and 2020.

**Measurements:** Annual overdose death rate, proportion of overdose deaths involving specific substances, and decedent demographics (age, sex, race, and ethnicity).

**Results:** We find emerging increases in annual opioid-related overdose death rates in Alaska (55.3% [*P=*0.020]), Colorado (80.2% [*P<*0.001]), Indiana (40.1% [*P=*0.038]), North Carolina (30.5% [*P<*0.001]), and Rhode Island (29.6% [*P=*0.011]). Decreased heroin-involved overdose deaths were emerging in Alaska (−49.5% [*P=*0.001]) and Indiana (−58.8% [*P<*0.001]), and continuing in Colorado (−33.3% [*P<*0.001]), Connecticut (−48.2% [*P<*0.001]), Massachusetts (39.9% [*P<*0.001]), and North Carolina (−34.8% [*P<*0.001]). Increases in synthetic opioid presence were emerging in Alaska (136.5% [*P=*0.019]) and Indiana (27.6% [*P<*0.001]), and continuing in Colorado (44.4% [*P<*0.001]), Connecticut (3.6% [*P<*0.05]), and North Carolina (14.6% [*P<*0.001]). We find emerging increases in the proportion of male decedents in Colorado (15.2% [*P=*0.008]) and Indiana (12.0% [*P=*0.013]).

**Limitations:** Delays from state-specific death certification processes resulted in varying analysis periods across states.

**Conclusion:** These findings highlight emerging changes in opioid overdose dynamics across different states, which can inform state-specific public health interventions.

## Introduction

Since COVID-19 erupted in the United States, experts warned that the pandemic’s strain on healthcare systems, economic stability, and social support structures would threaten many vulnerable individuals’ physical and mental well-being.^1,2^ Concurrently, the opioid overdose epidemic has continued to evolve, becoming more fatal since the onset of COVID-19.

Initial estimates indicate increases in opioid overdose admissions and deaths across emergency departments in San Francisco through April 18, 2020^3^ and Indianapolis through July 24, 2020.^4^ Recent provisional estimates from the United States Centers for Disease Control and Prevention indicate that opioid overdose deaths have increased through October 2020,^5^ with May 2020 being the deadliest month.^6^ These estimates, along with recent increases in overdose-related cardiac arrests nationwide,^7^ corroborate reports around the country which link increasing opioid overdose trends to COVID-19.^8^

This study extends previous analyses by: a) characterizing shifting trends in opioid overdose deaths by sex, age, race, and substances present; b) distinguishing between *emerging* trends (i.e., change from 2019-2020 was non-existent from 2018-2019) and *continuing* trends (i.e., change from 2019-2020 existed from 2018-2019); and c) analyzing confirmed state-level mortality data rather than provisional data.

## Methods

We submitted data requests to all 50 states and District of Columbia for vital statistics data from 2018-2020. Our data requests were completed for 12 states. Of those 12 states, we only included in our analysis nine states (Alaska, Colorado, Connecticut, Indiana, Massachusetts, North Carolina, Rhode Island, Utah, and Wyoming) that were able to provide vital statistics data extending beyond March 2020. [*We are finalizing the data request process with the vital statistics departments in Virginia and Nevada and plan to include these data after the first round of review if the data are received by that time*]. States not included in our analysis have pending data requests or do not report 2020 data. This study was deemed exempt from review by Mass General Brigham’s institutional review board.

We compared data from 2020 vs. 2019 to assess whether any changes in opioid overdose trends and decedent characteristics have occurred since the onset of COVID-19. Then, we compared data from 2019 vs. 2018 to confirm whether significant changes in 2020 vs. 2019 were *emerging* trends (i.e., no significant difference in 2019 vs. 2018) or *continuing* pre-existing trends (i.e., a significant difference in 2019 vs. 2018 in the same direction).

We defined separate analysis periods for each state beginning on March 13 (i.e., the day that COVID-19 was announced as a national emergency in the United States) and lasting until the latest reliable date of analysis as estimated in part by the state’s vital statistics experts. For Indiana, data were only available on a monthly basis and thus, we defined its analysis period from March 1 through June 30. These analysis periods differed across each state due to variation in each state’s COVID-19 response and death certification processes.^21^

For each state and year-over-year comparison, we compared the annual opioid overdose death rates per 100,000 people, opioid substances involved (i.e., ICD-10 codes T40.1, T40.2, T40.4, and T40.6), other psychoactive substances involved (i.e., ICD-10 codes T40.5, T42.4, and T43.6), and decedent demographics (i.e., age, sex, and race) from the analysis period in 2020 vs. 2019 and 2019 vs. 2018. For Alaska, Massachusetts, and Utah, the final and supporting causes of death were available in text format, where specific substances could be extracted. By analyzing the text data, we compared the mentions of seven categories of substances: fentanyl (and its analogues), prescription opioids (including hydrocodone, hydromorphone, oxycodone, oxymorphone, codeine, dihydrocodeine, levorphanol, and tramadol), benzodiazepines, amphetamines (including methamphetamine), and alcohol.

In all states, we compared the mean annual opioid overdose death rates across different years using the bootstrap two-sample t-test and the ICD-10 codes and decedent demographics using Pearson’s Chi-squared test. For Alaska, Massachusetts, and Utah, we also applied the Pearson’s Chi-squared test to assess for changes in the presence of substances derived from final and supporting causes of death text data. To determine which categories were driving significant differences in age and race, we performed post-hoc Chi-squared analyses on expected residuals^22^ using the Benjamini-Hochberg p-value correction for multiple comparisons.^23^ All data analyses were performed using R version 4.1.0.

## Results

### Overdose Death Rates

Our results comparing annual overdose deaths per 100,000 people and overdose death by substance are summarized in Table 1. We also illustrate the change from year to year in annual overdose deaths per 100,000 people in Figure 1. Comparing 2020 to 2019 in each state’s analysis period, the annual overdose death rate per 100,000 people has increased in Alaska (13.85 vs. 8.92, *P=*0.020), Colorado (18.24 vs.10.12, *P<*0.001), Indiana (25.98 vs. 18.54, *P=*0.038), North Carolina (22.79 vs. 17.47, *P<*0.001), and Rhode Island (30.39 vs.23.45, *P=*0.011). Compared to 2019 vs. 2018, these rising death rates are emerging in Alaska, Colorado, Indiana, North Carolina, and Rhode Island (i.e., they have *P*>0.05).

**Table 1.**
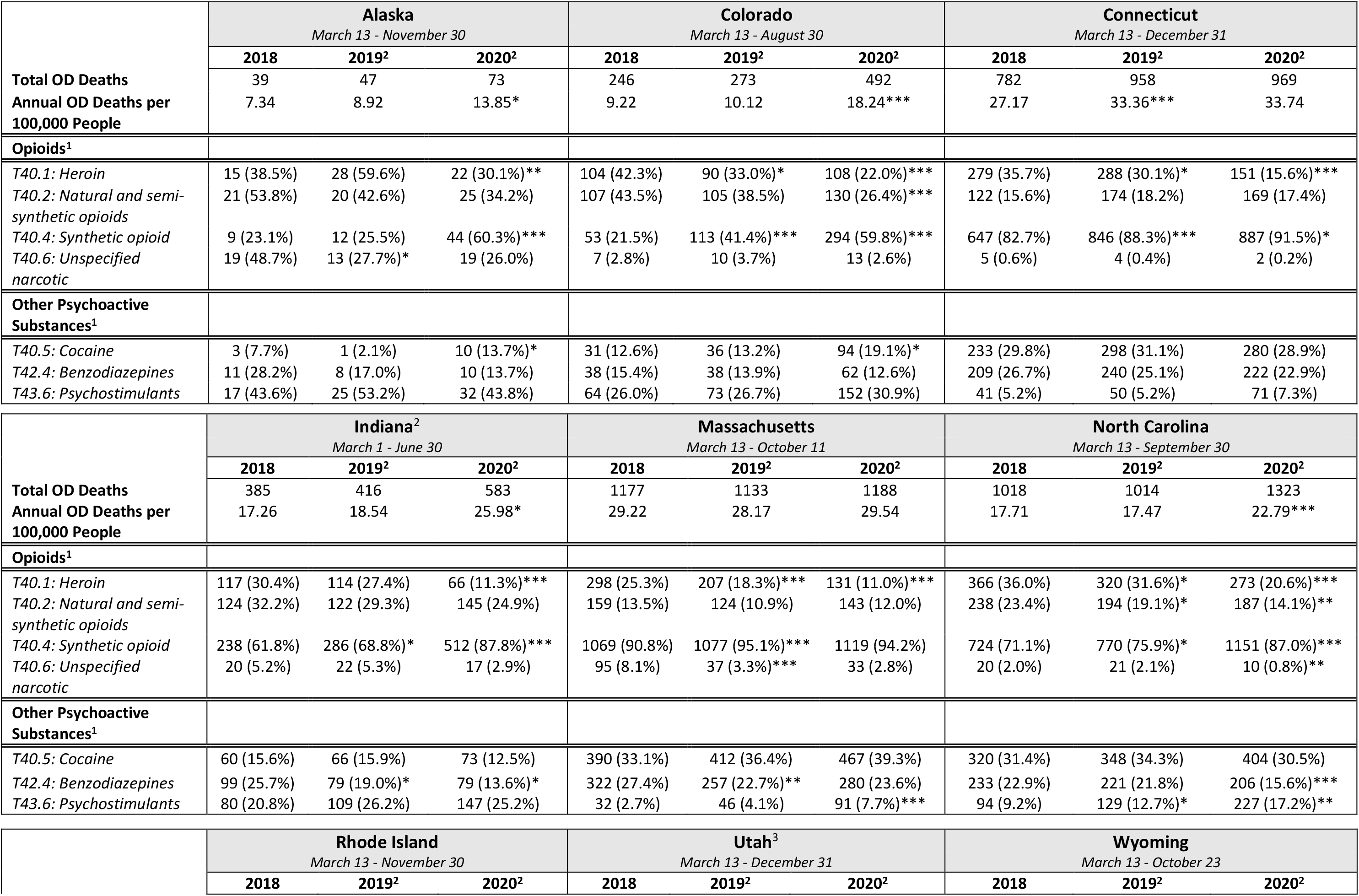
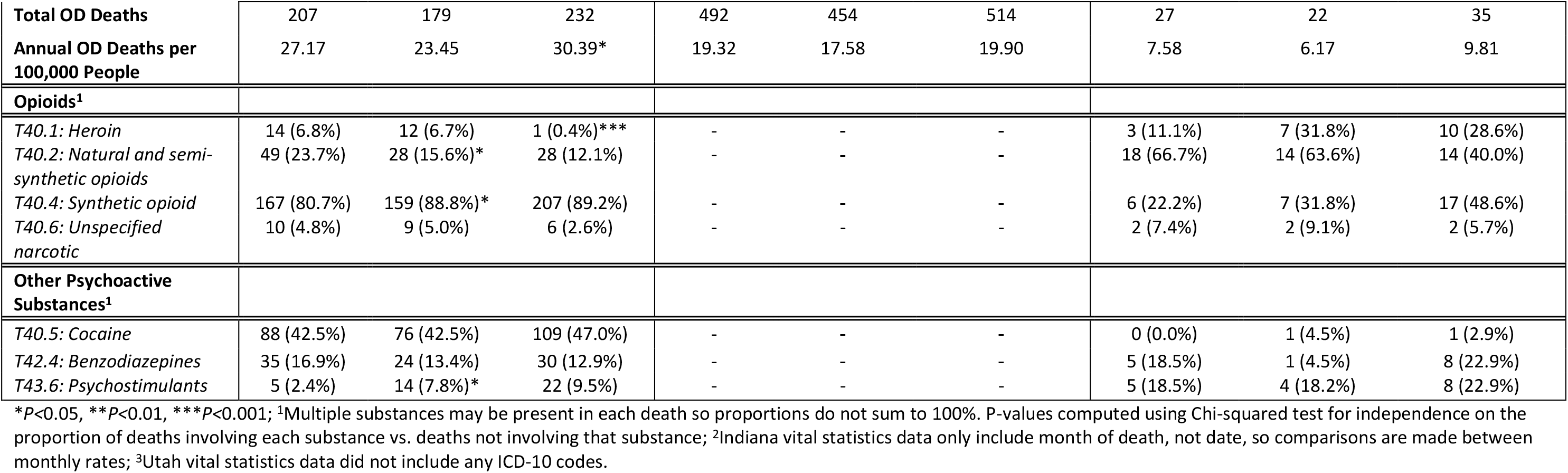
Analysis of opioid overdose death rates and substances present in 2018-2020.

**Figure 1.**
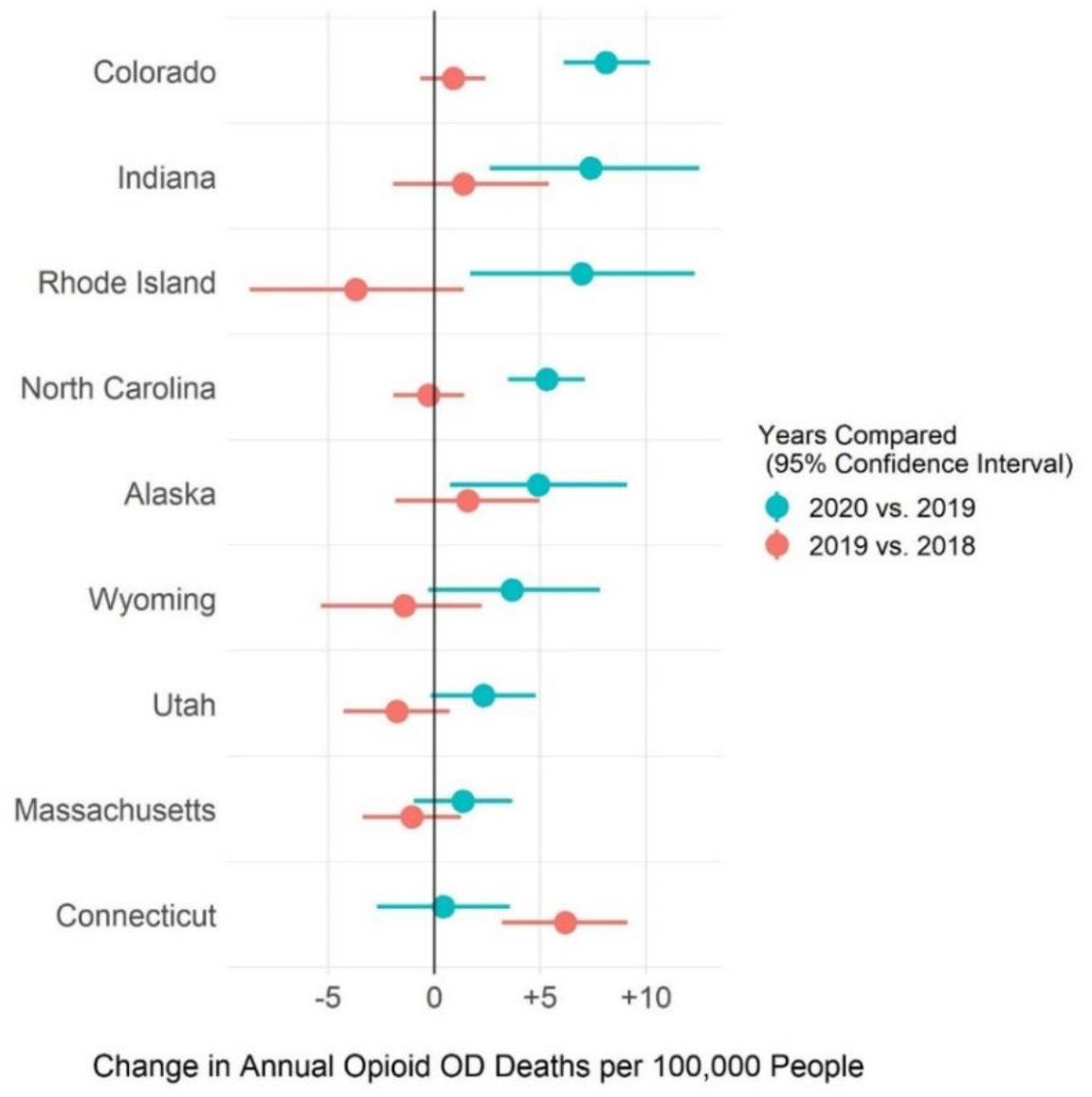
Changes in Annual Opioid Overdose Deaths per 100,000 People between 2020 vs. 2019 and 2019 vs. 2018 by State.

### Presence of Opioids

Trends have shifted regarding the type of opioids involved in these overdose deaths. Comparing 2020 vs. 2019, the proportion of heroin-involved opioid overdose deaths was significantly reduced in Alaska (30.1% vs. 59.6%, *P=*0.001), Colorado (22.0% vs. 33.0%, *P<*0.001), Connecticut (15.6% vs. 30.1%, *P<*0.001), Indiana (11.3% vs. 27.4%, *P<*0.001), Massachusetts (11.0% vs. 18.3%, *P<*0.001), North Carolina (20.6% vs. 31.6%, *P<*0.001), and Rhode Island (0.4% vs. 6.7%, *P<*0.001). Compared to 2019 vs. 2018, these trends were emerging in Alaska, Indiana, and Rhode Island, and continuing previous trends in Colorado (33.0% vs. 42.3%, *P=*0.028), Connecticut (30.1% vs. 35.7%, *P=*0.013), Massachusetts (18.3% vs. 25.3%, *P<*0.001), and North Carolina (31.6% vs. 36.0%, *P=*0.036).

Comparing 2020 to 2019, the proportion of natural and semi-synthetic opioid-involved overdose deaths was significantly reduced in Colorado (26.4% vs. 38.5%, *P<*0.001) and North Carolina (14.1% vs. 19.1%, *P=*0.001). Compared to 2019 vs. 2018, the shift in Colorado was emerging and the shift in North Carolina continues previous trends (19.1% vs. 23.4%, *P=*0.019).

Since the onset of COVID-19 (i.e., 2020 vs. 2019), there has also been a significant increase in the proportion of synthetic opioids among opioid-related overdose deaths in Alaska (60.3% vs. 25.5%, *P<*0.001), Colorado (59.8% vs. 41.4%, *P<*0.001), Connecticut (91.5% vs. 88.3%, *P=*0.019), Indiana (87.8% vs. 68.8%, *P<*0.001), and North Carolina (87.0% vs. 75.9%, *P<*0.001). Compared to 2019 vs. 2018, these increases were emerging in Alaska and continuing past trends in Colorado (41.4% vs. 21.5%, *P<*0.001), Connecticut (88.3% vs. 82.7%, *P<*0.001), Indiana (68.8% vs. 61.8%, *P=*0.039), and North Carolina (75.9% vs. 71.1%, *P=*0.014).

### Presence of Other Psychoactive Substances

Trends have also shifted with regard to the presence of non-opioid psychoactive substances. Comparing 2020 vs. 2019, there has been a significant increase in the proportion of opioid-related overdose deaths involving cocaine in Alaska (13.7% vs. 2.1%, *P=*0.032) and Colorado (19.1% vs. 13.2%, *P=*0.037), and psychostimulants in Massachusetts (7.7% vs. 4.1%, *P<*0.001) and North Carolina (17.2% vs. 12.7%, *P=*0.003). Compared to 2019 vs. 2018, these shifts were emerging in Alaska, Colorado, and Massachusetts and continuing previous trends in North Carolina (12.7% vs. 9.2%, *P=*0.012). 2020 vs. 2019 also brought reductions in the proportion of opioid-related overdose deaths involving benzodiazepines in Indiana (13.6% vs. 19.0%, *P=*0.020) and North Carolina (15.6% vs. 21.8%, *P<*0.001).

### Final and Supporting Causes of Death

Our results comparing text descriptions of final and supporting causes of death are presented in Table 2. The proportion of deaths involving fentanyl and fentanyl analogs increased in Alaska (58.9% vs. 12.8%, *P<*0.001) and Utah (21.0% vs. 8.8%, *P<*0.001). These trends are emerging in both states compared to 2019 vs. 2018. In 2020 vs. 2019, there were also increases in the proportion of opioid-related overdose deaths involving amphetamines (7.6% vs. 4.2%, *P<*0.001) and alcohol (27.3% vs. 22.9%, *P=*0.014) in Massachusetts. Each of these trends were emerging compared to 2019 vs. 2018.

**Table 2.**
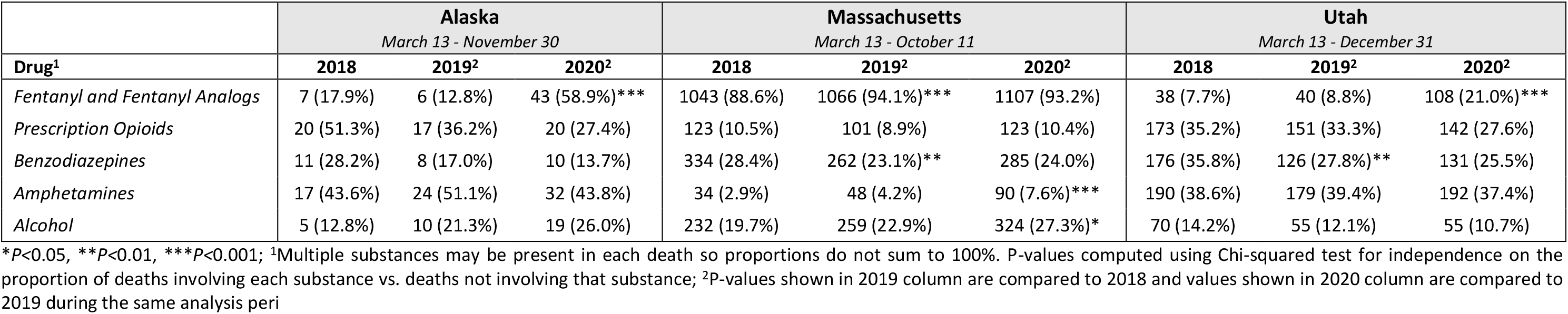
Analysis of official and supporting causes of death in Alaska, Massachusetts, and Utah.

### Decedent Demographics

Our analysis of decedent demographics is summarized in Supplementary Table S1. Comparing 2020 vs. 2019, Colorado (*P=*0.008) and Indiana (*P=*0.013) experienced significant shifts in the sex of decedents. These shifts were driven by an increase in the proportion of male decedents in both Colorado (70.5% vs. 61.2%, *P=*0.017) and Indiana (70.0% vs. 62.5%, *P=*0.026). Compared to 2019 vs. 2018, both shifts in decedent sex were emerging. North Carolina also witnessed a shift in decedent race (*P<*0.001) in 2020 vs. 2019. This shift was driven by a decrease in the proportion of Native American/Alaska Native/Other non-Hispanic decedents (0% vs. 2.0%, *P<*0.001) and White non-Hispanic decedents (75.5% vs. 81.1%, *P=*0.0044) as well as an increase in the proportion of Hispanic (4.4% vs. 2.6%, *P=*0.048) decedents and decedents of unknown race (4.7% vs. 0.8%, *P<*0.001). This shift in decedent race was emerging compared to 2019 vs. 2018. There were no significant shifts in age for any state.

## Discussion

The COVID-19 pandemic has complicated public health efforts to mitigate and control the ongoing opioid crisis. This analysis extends previous single-city and single-state reports on opioid overdose deaths since the onset of COVID-19. More importantly, this research highlights emerging and continuing trends in shifting demographics and substance use patterns among opioid overdose decedents across nine states during the pandemic.

News reports across the United States since the onset of COVID-19 have alluded to rising opioid overdose death rates in many states.^8^ While our results show increases in the annual overdose death rate across several states in our study, these increases were only significant in Alaska, Colorado, Indiana, North Carolina, and Rhode Island. Notably, each of these statistically significant increases were emerging trends in 2020. While it is challenging to disentangle the exact causes and further investigations are required, the rising overdose death rates in these states could potentially be attributed, in part, to COVID-19 related stress and isolation, limited access to in-person harm reduction services or treatment, as well as an increasing presence of fentanyl in the illicit drug supply.^9,10^ Nevertheless, it is plausible that each state in our analysis may have also seen a significant rise in overdose deaths related to COVID-19. However, these deaths may have been offset by other pre-existing opioid overdose trends or overdose mitigation strategies that emerged in anticipation of the pandemic’s impact. To this end, ascertaining the exact causes of changing trends, while important, is beyond the scope of our analysis.

In Colorado and Indiana, there have been statistically significant increases in the proportion of male decedents since the beginning of each state’s initial stay-at-home orders. Previous research has drawn associations between increased opioid overdose rates during times of increased financial hardship and unemployment, especially among men.^11,12^ These increases could potentially be related to financial impact of COVID-19, although additional analyses would be needed to confirm any causal relationship.

In every state we analyzed, there was an increasing proportion of Black non-Hispanic decedents and decreasing proportion of White non-Hispanic decedents since the start of the pandemic. To this end, only the shift in racial demographics in North Carolina was statistically significant, although it could not be attributed to the increase in Black non-Hispanic decedents. While further research is needed to identify the source of these shifts in racial demographics, these data might reflect the potentially compounding effects of COVID-19 on already recently increasing rates of synthetic opioid-related deaths among Black people.^13^

Across most states we analyzed, there was an increase in synthetic opioid-related deaths (e.g., fentanyl). Notably, this increase was only emergent in Alaska and was otherwise a reflection of ongoing trends in the other states. Simultaneously, most states also experienced a reduction in the proportion of heroin-involved overdose deaths. National-level analyses have shown steady rates of heroin presence in opioid overdose deaths through 2018,^14^ so these results may indicate the beginning of a greater trend in the reduction of heroin presence overall. Our analysis of final and supporting causes of death underscore these findings, while also revealing an emerging increase in amphetamine- and alcohol-involved overdose deaths in Massachusetts. The increased presence of these non-opioid substances may reflect signals of increased alcohol and stimulant use during the pandemic as reported in various cities across the United States^15,16^, although these increasing trends were pre-existing at a national level.^17^

Our study was limited by several data challenges.^18^ Inconsistencies in data formatting complicated our analysis. For example, some states could not provide the exact dates of each death (e.g., only the month and year was available), leading to less granular data and a loss of statistical power in estimating overdose death rates from aggregation. Additionally, delays from the death certification process in each state resulted in varying analysis periods for each state, and in some cases, prevented us from performing any analysis at all due to a lack of 2020 data. Notably, we received data from Maryland, Mississippi, and Ohio but these states were excluded from our analysis because they could only provide data until the end of 2019. Given the gravity of the opioid crisis in the United States, these limitations affect all analyses which utilize fatal opioid overdose data and highlight the need to collectively improve data collection and reporting infrastructure.

## Conclusions

This research analyzed changing trends in opioid overdose decedent characteristics since the onset of the COVID-19 pandemic across seven states. To this end, various health policy researchers and organizations such as the Centers for Disease Control and Prevention^19^ and American Medical Association^20^ have provided recommendations to mitigate the effects of the COVID-19 pandemic on the opioid overdose crisis. Our findings highlight emerging trends which can further inform each state’s response to these intersecting crises. More broadly, the trends uncovered in this analysis signal potential trends that may emerge for states with data that remain to be analyzed.

## Data Availability

Data can be requested by contacting the corresponding author.

## Online Supplementary Materials for

**Supplementary Table S1.**
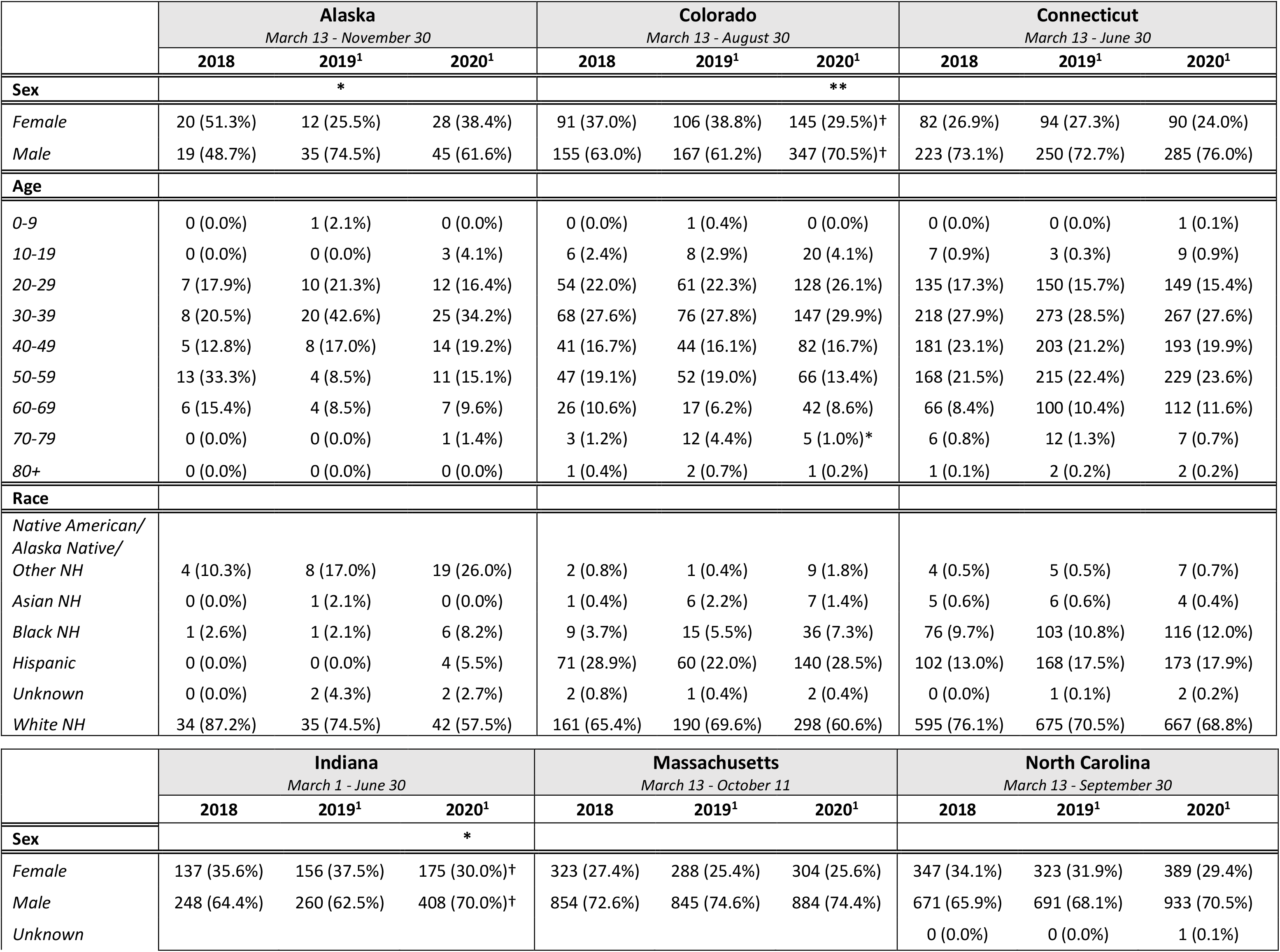
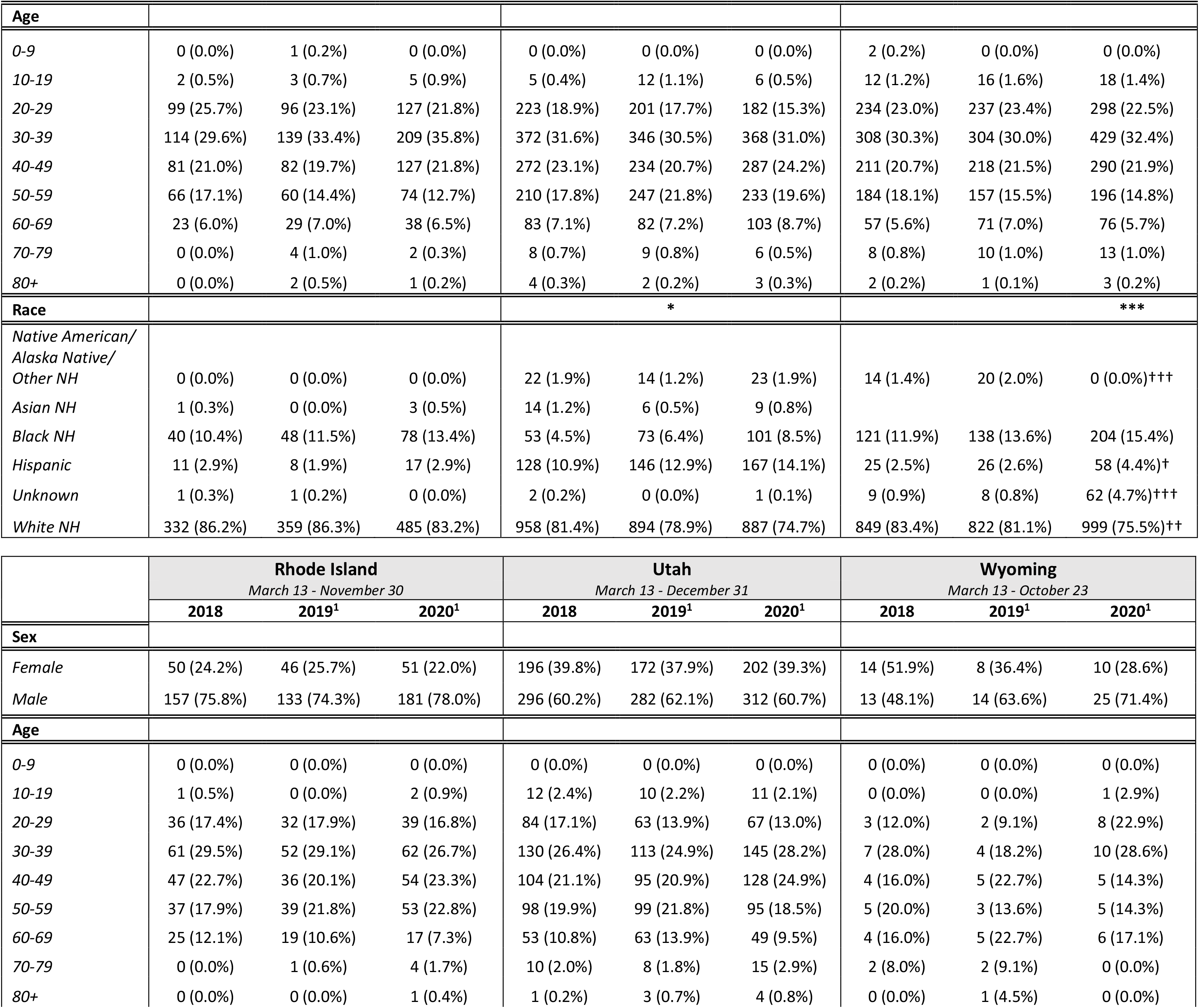
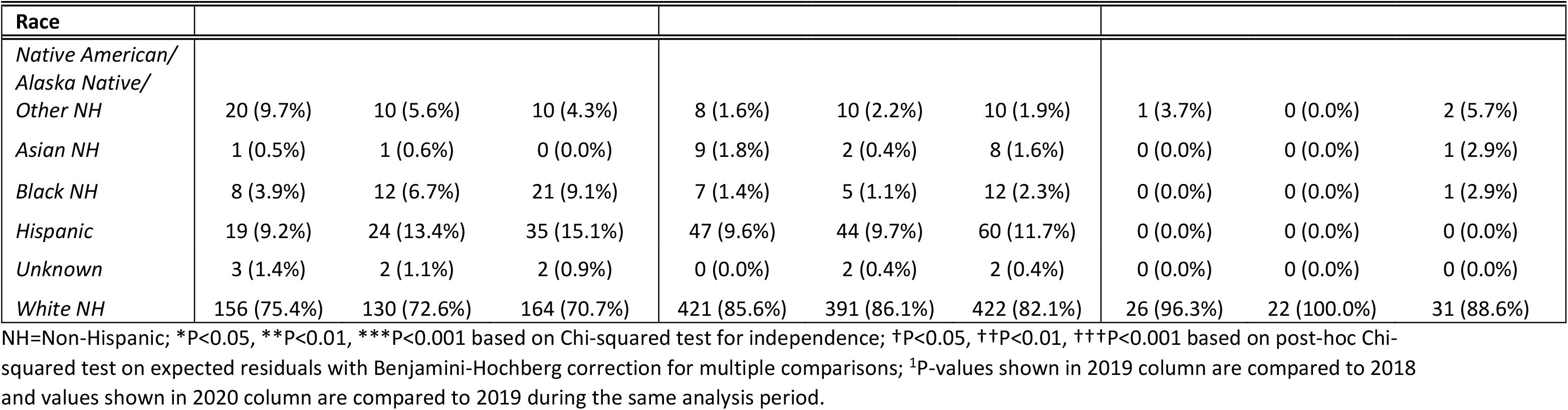
Analysis of demographics among opioid-related overdose decedents, 2018-2020

